# Trends in cognitive function before and after diabetes onset in China

**DOI:** 10.1101/2023.07.02.23292154

**Authors:** Sijia Zhu, Qingmei Chen, Jing Shang, Jianian Hua

**Author notes:** <u>Correspondence:</u> Jianian Hua. These authors contributed equally (Sijia Zhu, Qingmei Chen).

## Abstract

**Background:** Individuals with prevalent diabetes were reported to have higher risk of dementia and lower cognitive function. However, the trends of cognitive function before diabetes and in the years following diabetes onset remain unclear.

**Methods and Findings:** This study included 12422 participants aged ≥45 years without baseline diabetes from the China Health and Retirement Longitudinal Study (CHARLS). Cognitive function was assessed at baseline (Wave 1, 2011), and at least one time from Wave 2 (2013) to Wave 4 (2018). During the 7-year follow-up, 1207 (9.7%, 59.1 ± 8.6 years, 39.9% males) participants developed new-onset diabetes. The cognitive function of both the without-diabetes group and the diabetes group declined annually during the follow-up. The annual decline rate of the diabetes group before diabetes onset was similar to that of the without-diabetes group during the follow-up. After diabetes onset, participants experienced accelerated rates of cognitive decline in global cognition (β, −0.023 SD/year; 95% CI, −0.043 to −0.004) and visuospatial abilities (−0.036 SD/year; −0.061 to −0.011), but not in orientation abilities (0.001 SD/year; −0.018 to 0.020). We also observed a tendency that episodic memory (−0.018 SD/year; −0.041 to 0.004) and attention and calculation abilities (−0.017 SD/year; −0.037 to 0.003) declined faster after new-onset diabetes, although the results did not meet our threshold of significance. In subgroup analysis, compared with those who developed diabetes between 45–54 years old, those developing diabetes older showed similar increments in cognitive decline rate after diabetes.

**Conclusions:** Individuals experienced faster rate of cognitive decline after diabetes onset, but not during the pre-diabetes period. Age did not modify the effect of diabetes on future cognitive decline. Future studies are needed to learn the mechanisms of cognitive decline in a few years after new-onset diabetes.

## 1 Introduction

Dementia affects around 50 million people worldwide. The number is predicted to increase to 152 *million* by 2050 (1). Due to the lack of effective treatments, modifiable risk factors have drawn special attention for dementia prevention. Diabetes is a key modifiable risk factor for dementia (2). Hyperglycaemia contributes to dementia through both Alzheimer’s-Disease-related and vascular disease-related pathology, while the latter might be a major cause (3, 4).

Diabetes increased the risk of follow-up dementia (5). Meanwhile, those with known diabetes had lower cognitive function cross-sectionally and faster cognitive decline longitudinally (6, 7). Most studies only interviewed diabetes status at baseline, reflecting the cognitive trends among participants with prevalent diabetes. However, hyperglycemia might develop several years before the cognitive assessment. Cognitive function before or in the short period after diabetes onset is unknown. Considering that prediabetes could also affect cognitive health, the cognitive deterioration might begin even before diabetes (8). Dementia is a syndrome characterized by declines in cognitive function, understanding the cognitive deterioration before and in the short period after new-onset diabetes could help unravel the pathophysiology of diabetes on the brain and the prevention and management of dementia in diabetic individuals.

Few studies have indeed reported cognitive trends after (9, 10, 11, 12), or before and after new-onset diabetes (13, 14, 15). However, several knowledge gaps remain unaddressed. Firstly, the study population was relatively older in the previous two studies which calculated the cognitive trends before and after diabetes (14, 15). A recent article published in the JAMA reported younger age at onset of diabetes was associated with higher risk of subsequent dementia (16). Therefore, whether individuals with lower mean age exhibited different trajectories of cognitive function is not clear. Second, to our knowledge, cognitive trajectories before and after diabetes onset have not been investigated in the Chinese population, who have different educational and economic levels. This might contribute to different cognitive reserve and different patterns of cognitive trajectories (8). Third, cognitive decline is not a unitary process and may have disparate effects in different cognitive domains. Previous studies achieved mixed results in domain-specific cognitive trends. For example, some reported accelerated declines in memory function after diabetes onset (10, 11, 14), while some reported that memory function did not deteriorate after diabetes onset (14, 15).

Therefore, using data from the China Health and Retirement Longitudinal Study (CHARLS), we explore the temporal pattern of declines in cognitive function before and after diabetes onset among middle-aged and elderly Chinese population (≥ 45 years old).

## 2 Methods

### 2.1 Study participants

The current study used data from Wave 1 to Wave 4 of the CHARLS, a nationally representative and community-based cohort study in China (17). At baseline (Wave 1, 2011), 17,708 participants were recruited from 150 national units distributed in 28 provinces of China. The baseline survey was performed between June 2011 and March 2012. Three follow-up visits (Wave 2, 2013; Wave 3, 2015; and Wave 4, 2018) were conducted after baseline.

The flowchart for sample selection is shown in Figure 1. Out of the 16,040 participants who finished cognitive assessments at baseline, 1196 were excluded for the following reasons: under 45 years at baseline (n == 352), history of brain injuries or psychiatric disorders (n == 392), having a prior history of stroke (n == 423), or suffering from memory-related diseases (n == 206) which included Alzheimer’s disease, brain atrophy, and Parkinson disease. Subsequently, 2372 of the remaining 14,844 individuals were further excluded due to diabetes history at baseline (n == 423), lack of baseline covariates (n == 44), or not receiving at least one cognitive assessment during the follow-up from wave 2 to wave 4 (n == 1377). Ultimately, 12422 individuals consisting of 1207 participants with new-onset diabetes and 11,215 participants without diabetes were enrolled in the main analysis.

**Fig 1.**
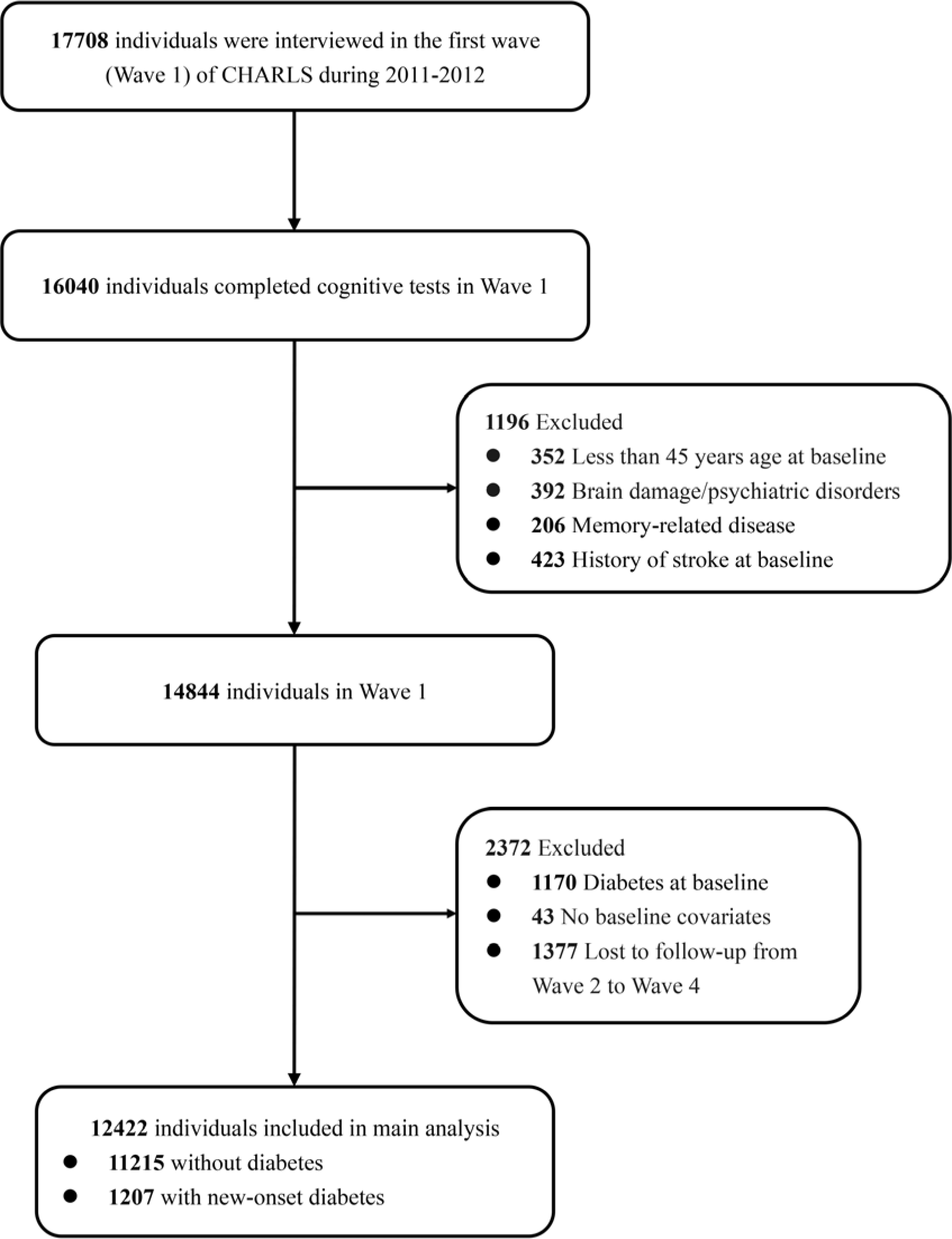
Flow chart of sample selection.

### 2.2 Assessment of cognitive function

Cognitive function was evaluated in each wave and included four domains: episodic memory, visuospatial abilities, orientation, and attention and calculation. The episodic memory was tested by word recall and its total score was composed of the correct number of immediate and delayed (5 minutes) word recalls. The score ranged from 0 to 10. Visuospatial abilities were evaluated by replicating two overlapping pentagons in a drawing. Participants who completed the task successfully received 1 point. The orientation abilities were assessed by asking participants to identify the date (year, month, day), day of the week, and season (score ranging, 0–5). The attention and calculation abilities were measured by repeatedly subtracting 7 from 100 for five times (score ranging, 0–5). The reliability and validity of these tests have been well documented in the previous investigations (17, 18). Z scores were created to allow direct comparisons across different cognitive domains by using the mean and SD of the scores at the baseline. The global cognitive z score was estimated by summing the z scores of the above four tests and then standardizing it to baseline in the same manner (19). Thus, a Z score of 1 would describe cognitive performance that is 1 SD above the mean score at the baseline.

### 2.3 Assessment of diabetes onset

Diabetes was defined as self-reported physician’s diagnosis, antidiabetic medication use, or hemoglobin A_1c_ (HbA_1c_) ≥6.5%. Information on self-reported diagnosis and medication use was available in all four waves. The HbA_1c_ level was measured in Wave 1 and Wave 3. For some participants, the time of diabetes onset was recorded through interviews (“When was diabetes first diagnosed or known?”). For other participants, the time of diabetes onset was recorded as the midpoint between the last time without diabetes and the first time with diabetes.

### 2.4 Covariates

Information on sociodemographic factors, lifestyle behaviors, and health-related variables was collected. Our analysis adjusted baseline covariates associated with diabetes and cognitive function, including age, sex, education, marital status, residential area, smoking, drinking, the number of instrumental activities of daily living (IADLs) for which the participant needs help, hypertension, high total cholesterol, cancer, lung diseases, heart problems, and depressive symptoms. Details of covariates are available in Text S2 in Appendix S1 (17).

### 2.5 Statistical analysis

Baseline characteristics were compared between participants who developed diabetes during the follow-up and those without diabetes till the end of the follow-up. We constructed a linear mixed model to analyze the longitudinal dataset with repeated measurements (20, 21). We fitted fixed effects for intercept, diabetes (yes or no), time (years since baseline), ‘diabetes*time’ interaction, ‘diabetes*diabetes status*time after diabetes’ interaction, and all the covariates. We fitted random effects for intercept and slope (time since baseline and time after diabetes). The estimate for the parameters could reflect the baseline cognitive difference between the diabetes group and the control group, the difference in the cognitive change rate (slope) between the diabetes group during the pre-diabetes period and the control group during the whole follow-up period, and whether the cognitive slope changed after diabetes onset compared with the cognitive slope before diabetes. Test S2 and Fig S1–S3 in Appendix S1 demonstrate the conceptual model and more detailed descriptions of statistical methods. As an example, in Fig 2, we show the cognitive trends of two fictitious persons (one with and the other without diabetes) with the most common baseline characteristics (19).

**Fig 2.**
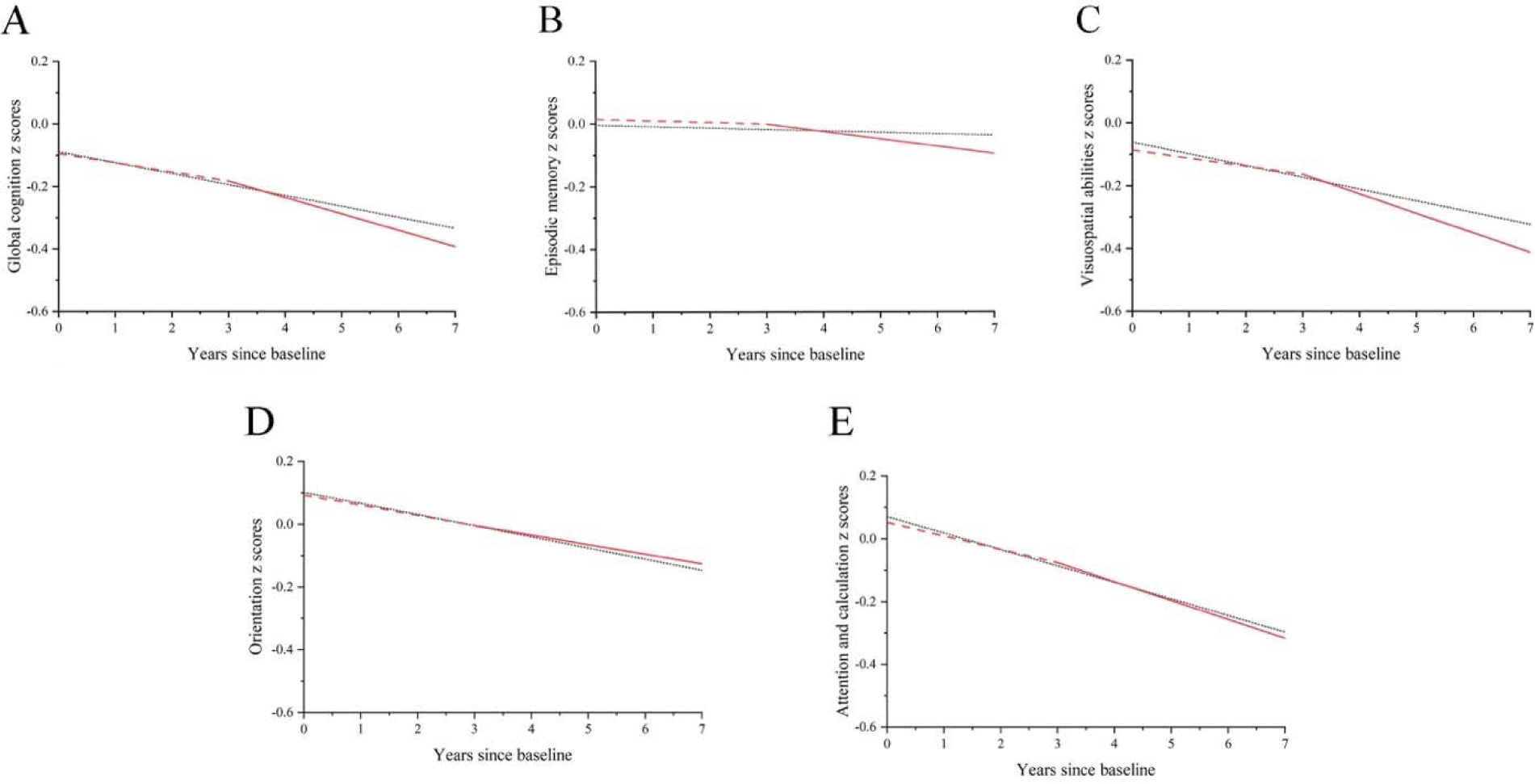
Predicted changes in cognitive z scores during the follow-up. The black dotted curves represent participants without diabetes. The red dashed curves represent the pre-diabetes cognitive trajectories among participants who developed diabetes during the follow-up. The red solid curves represent trajectories after new-onset diabetes. The predicted values of cognitive score are calculated for a 60-year-old female. Her education level is primary school. She lives in a rural area and is married but without current smoking, current drinking, hypertension, dyslipidaemia, cancer, lung diseases, heart problems, or depression. Her IADL score is zero. She developed new-onset diabetes at the end of the third year.

To explore the association between age at diabetes onset and the extent of cognitive decline, we calculated the trends among participants who developed diabetes between 45–55, 55–65, and 65– 75 years old, separately. The classification was based on the sample size of our diabetes participants across ages, and that a previous article reported that compared with those with diabetes onset at age 70, those with diabetes onset earlier had a higher risk of subsequent dementia (16). Using interaction terms, we tested whether the age group modified the intercept and slope of the cognitive trajectories (22). We further explore the interaction effect of the potential risk factors for cognitive decline, including sex, cognitive reserve (education and baseline cognitive scores), residential areas, and medication after diabetes. While exploring risk factors, we restricted our analysis only to those with diabetes due to the following reasons. First, the current study primarily focused on changes in cognitive slope after diabetes onset. Second, removing the control group could achieve a better model fit and make the results more interpretable.

To evaluate the stability of our main results, several sensitivity analyses were performed. a) Adding ‘diabetes*diabetes status’ as a fixed effect in the linear mixed model. This reflected whether the cognitive function declined acutely at diabetes onset (23, 24). A detailed description of this variable is provided in Figure S3. b) Adding BMI as a covariate. Since few participants had BMI data in Wave 1, measures of BMI in Wave 2 were used as the estimates in Wave 1 in this analysis. c) Excluding participants who suffered a stroke during the follow-up. d) Restricting participants to those who had complete cognitive data in all four waves.

All statistical analyses were performed using SAS version 9.4 (SAS Institute Inc., Cary, NC, USA). All *p* values were two-sided, with 0.05 being the threshold for statistical significance.

## 3 Results

### 3.1 Study participants

Over the 7-year follow-up, 1207 participants (9.7%, 59.1 ± 8.6 years at baseline, 39.9% males) developed new-onset diabetes. The number and diagnosis time of diabetes are disclosed in Table S1 in Appendix S1. Table 1 presents the baseline characteristics according to occurrence of diabetes. The mean ± age of diabetes-free individuals was 58.4 ± 9.3 years, while individuals with diabetes was 59.1 ± 8.6 years. Among the individuals with diabetes, 70.4% had not completed middle school. Moreover, the diabetes group tended to be female, had a higher number of IADLs, lower proportion of being unmarried, current smoking, and current drinking, and were more likely to have hypertension, dyslipidaemia, cancer, heart problems, and depression at baseline. Compared with the control group, participants who experienced diabetes showed lower baseline scores in domains of orientation and attention and calculation. However, the difference became insignificant once age and sex were taken into account.

**Table 1.**
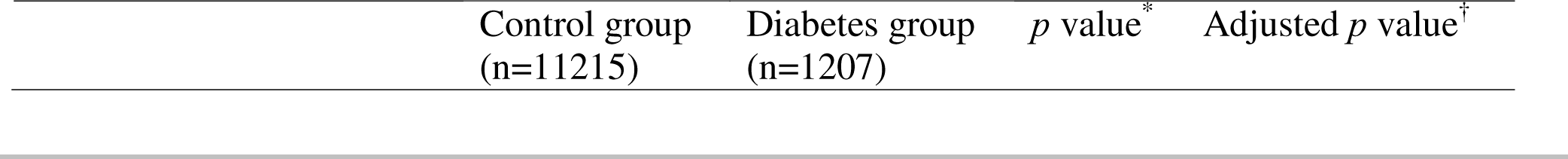
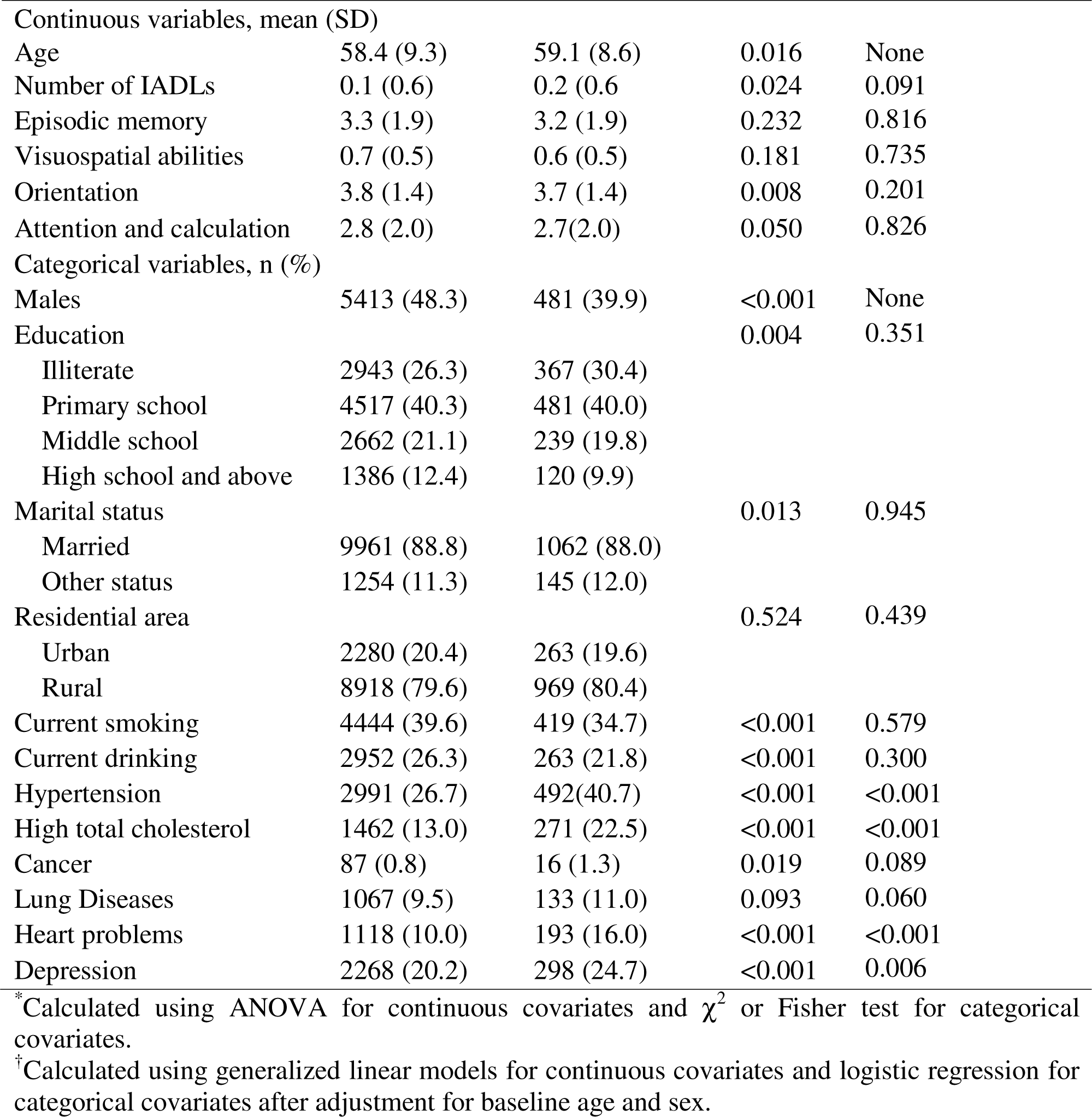
Baseline participant characteristics according to diabetes onset

| Control group<br>(n=11215) | Diabetes group<br>(n=1207) | *p* value<sup>*</sup> | Adjusted *p* value<sup>†</sup> |
| --- | --- | --- | --- |

|  |  |  |  |  |
| --- | --- | --- | --- | --- |
| Continuous variables, mean (SD) |  |  |  |  |
| Age | 58.4 (9.3) | 59.1 (8.6) | 0.016 | None |
| Number of IADLs | 0.1 (0.6) | 0.2 (0.6) | 0.024 | 0.091 |
| Episodic memory | 3.3 (1.9) | 3.2 (1.9) | 0.232 | 0.816 |
| Visuospatial abilities | 0.7 (0.5) | 0.6 (0.5) | 0.181 | 0.735 |
| Orientation | 3.8 (1.4) | 3.7 (1.4) | 0.008 | 0.201 |
| Attention and calculation | 2.8 (2.0) | 2.7(2.0) | 0.050 | 0.826 |
| Categorical variables, n (%) |  |  |  |  |
| Males | 5413 (48.3) | 481 (39.9) | <0.001 | None |
| Education |  |  | 0.004 | 0.351 |
| Illiterate | 2943 (26.3) | 367 (30.4) |  |  |
| Primary school | 4517 (40.3) | 481 (40.0) |  |  |
| Middle school | 2662 (21.1) | 239 (19.8) |  |  |
| High school and above | 1386 (12.4) | 120 (9.9) |  |  |
| Marital status |  |  | 0.013 | 0.945 |
| Married | 9961 (88.8) | 1062 (88.0) |  |  |
| Other status | 1254 (11.3) | 145 (12.0) |  |  |
| Residential area |  |  | 0.524 | 0.439 |
| Urban | 2280 (20.4) | 263 (19.6) |  |  |
| Rural | 8918 (79.6) | 969 (80.4) |  |  |
| Current smoking | 4444 (39.6) | 419 (34.7) | <0.001 | 0.579 |
| Current drinking | 2952 (26.3) | 263 (21.8) | <0.001 | 0.300 |
| Hypertension | 2991 (26.7) | 492(40.7) | <0.001 | <0.001 |
| High total cholesterol | 1462 (13.0) | 271 (22.5) | <0.001 | <0.001 |
| Cancer | 87 (0.8) | 16 (1.3) | 0.019 | 0.089 |
| Lung Diseases | 1067 (9.5) | 133 (11.0) | 0.093 | 0.060 |
| Heart problems | 1118 (10.0) | 193 (16.0) | <0.001 | <0.001 |
| Depression | 2268 (20.2) | 298 (24.7) | <0.001 | 0.006 |

\*Calculated using ANOVA for continuous covariates and $\chi^2$ or Fisher test for categorical covariates.
†Calculated using generalized linear models for continuous covariates and logistic regression for categorical covariates after adjustment for baseline age and sex.

From waves 1 to 4, the available cognition measurements were 11215, 9974, 9675, and 7678 in the without-diabetes group, and 1207, 1112, 1120, and 930 in the diabetes group, respectively (Appendix S1: Table S2). 1377 (11.1%) participants not attending any follow-up interviews were excluded. These lost-to-follow-up were older; had lower scores in all cognitive domains and a higher number of IADLs. Moreover, they were less likely to be married, live in rural areas, drink, or have high total cholesterol; had a higher percentage of hypertension, cancer, lung disease, lung diseases, heart problems, and depression (Appendix S1: Table S3).

### 3.2 Trajectories in cognitive scores

For those without diabetes, the cognitive scores declined annually due to cognitive aging, which is shown by the black dotted lines in Fig 2. No difference in baseline was observed between the diabetes group and the control group. The decline rates from baseline to diabetes onset of the diabetes group were not statistically different from that of the control group from baseline to the end of follow-up (Table 2, Table S4 in Appendix S1). The above results illustrated that there was no pre-diabetes cognitive disadvantage in our diabetes group.

**Table 2.** Difference in cognitive z scores over time among all participants^a^

|  | Baseline difference |  | Difference in slope before diabetes onset |  | Difference in slope after diabetes onset |  |
| --- | --- | --- | --- | --- | --- | --- |
| | $\beta$ (95% CI) | *p* | $\beta$ (95% CI) | *p* | $\beta$ (95% CI) | *p* |
| Global cognition | -0.006 (-0.055, 0.042) | 0.802 | 0.006 (-0.007, 0.019) | 0.376 | -0.023 (-0.043, -0.004) | 0.019 |
| Episodic memory | 0.018 (-0.039, 0.076) | 0.530 | 0.000 (-0.016, 0.015) | 0.951 | -0.018 (-0.041, 0.004) | 0.116 |
| Visuospatial abilities | -0.025 (-0.082, 0.033) | 0.400 | 0.011 (-0.005, 0.028) | 0.178 | -0.036 (-0.061, -0.011) | 0.004 |
| Orientation | -0.009 (-0.057, 0.039) | 0.713 | 0.003 (-0.009, 0.016) | 0.603 | 0.001 (-0.018, 0.020) | 0.894 |
| Attention and calculation | -0.019 (-0.074, 0.036) | 0.496 | 0.010 (-0.004, 0.024) | 0.174 | -0.017 (-0.037, 0.003) | 0.090 |

<sup>a</sup>Adjusted for baseline age, sex, education, marital status, residential area, current smoking, current drinking, hypertension, dyslipidaemia, cancer, lung diseases, heart problems, depression, and number of IADLs.

In the years after diabetes onset, scores in all cognitive domains declined over time (Fig 2, Table S4 in Appendix S1). For the control group, the global cognitive scores declined by a mean of 0.035 SD/year. The slope of global cognition scores after diabetes onset was steeper than it was before diabetes (β, −0.023 SD/year; 95% CI, −0.043 to −0.004; *p*=0.019; Table 2 and Fig 2), roughly 68% faster (95% CI, 44% to 90%). Similar results were found in visuospatial abilities scores (β, −0.036 SD/year; 95% CI, −0.061 to −0.011; *p*=0.004). The decline rate of visuospatial abilities scores after diabetes onset was about 96% faster (95% CI, 68% to 125%) than that of the control group. We observed a trend that the decline rate of episodic memory scores (β, −0.018 SD/year; 95% CI, −0.041 to 0.004; *p*=0.116) and scores of attention and calculation abilities (β, −0.017 SD/year; 95% CI, −0.037 to 0.003; *p* =0.090) accelerated after diabetes. However, the difference in slope is not statistically significant. The decline rate of orientation abilities scores after diabetes was similar to the rate before diabetes (β, 0.001 SD/year; 95% CI, −0.018 to 0.020; *p*=0.894).

### 3.3 Risk factors for cognitive decline

There were 275 participants developed diabetes between 45–54 years old, and 478 between 55– 64 years old, and 352 between 65-74 years old, respectively. Difference in cognitive slope after diabetes onset compared with the cognitive slope before diabetes are shown in Table 3. Compared with participants who developed diabetes between 45–54 years old, those developing diabetes between 55–64 years old or between 65-74 years old did not experience significant increment in accelerated cognitive decline in the years following diabetes onset. Sex and residential area did not modify the effect of diabetes on accelerated cognitive decline after diabetes. Compared with participants who had not finished middle school, those who finished middle school or high school showed slower after-diabetes-cognitive-decline in episodic memory scores, but not in other cognitive domains. Participants in higher baseline cognitive tertiles had significantly slower decline in all cognitive domains after diabetes, compared with participants in the lowest cognitive tertile. Compared with participants who did not use antidiabetic medication after diabetes, those who reported using antidiabetic medication exhibited faster decline in global cognition after diabetes (Appendix S1: Table S5–S9).

**Table 3.** Changes in cognitive z scores after diabetes according to age at diabetes onset^a^

|  | 45–54 y (n=275) | 55–64 y (n=478) | 65–74 y (n=352) |
| --- | --- | --- | --- |
| Global cognition |  |  |  |
| β (95% CI) | -0.028 (-0.068, 0.011) | -0.031 (-0.062, -0.001) | -0.030 (-0.067, 0.008) |
| p for interaction<sup>b</sup> | Ref. | 0.701 | 0.996 |
| Episodic memory |  |  |  |
| β (95% CI) | -0.014 (-0.063, 0.034) | -0.049 (-0.085, -0.013) | -0.021 (-0.064, 0.021) |
| p for interaction | Ref. | 0.576 | 0.903 |
| Visuospatial abilities |  |  |  |
| β (95% CI) | -0.037 (-0.086, 0.013) | -0.046 (-0.085, -0.007) | -0.028 (-0.076, 0.019) |
| p for interaction | Ref. | 0.651 | 0.491 |
| Orientation |  |  |  |
| β (95% CI) | 0.007 (-0.034, 0.048) | 0.019 (-0.012, 0.05) | -0.018 (-0.055, 0.02) |
| p for interaction | Ref. | 0.614 | 0.211 |
| Attention and calculation |  |  |  |
| β (95% CI) | -0.029 (-0.079, 0.020) | -0.015 (-0.048, 0.018) | -0.015 (-0.052, 0.022) |
| p for interaction | Ref. | 0.170 | 0.460 |

<sup>a</sup>Adjusted for baseline age, sex, education, marital status, residential area, current smoking, current drinking, hypertension, dyslipidaemia, cancer, lung diseases, heart problems, depression, and number of IADLs.
<sup>b</sup>The interaction term reflected whether age groups modified the effect of diabetes on the cognitive decline rate after diabetes onset.

### 3.4 Sensitivity analyses

We did not observe ‘acute cognitive decline’ at the time of diabetes onset (Appendix S1: Table S10). After adding BMI as a covariate, excluding participants who suffered a stroke during the follow-up, or restricting participants receiving cognitive measurments in all four waves, we achieved similar results. In all sensitivity analyses, the scores of global cognition and visuospatial abilities declined gradually before diabetes and showed a faster decline rate in the years after diabetes onset (Appendix S1: Table S11-S13).

## 4 Discussion

In this population-based study of Chinese adults aged ≥45 years, participants experienced faster rates of cognitive decline in global cognition and visuospatial abilities after diabetes onset, compared with the rate before diabetes. We also observed the tendency that episodic memory and attention and calculation scores declined faster after diabetes. For orientation abilities, the cognitive decline did not accelerate after diabetes. The rate of cognitive decline before diabetes onset among those with diabetes was similar with the rate of individuals free of diabetes.

The cognitive trajectory before and after new-onset diabetes was relatively unexplored, especially in Asia. Several longitudinal studies have assessed their participants’ cognitive function more than once and calculated the cognitive trends of those who developed new-onset diabetes during the follow-up period (9, 10, 11, 12, 13). However, they did not analyze whether the trends in cognition after diabetes onset differed from that before diabetes. To our knowledge, only 2 studies compared the cognitive trajectory before and after diabetes onset. A Chicago Health and Aging Project (CHAP) study reported faster post-diabetes declines in Mini-Mental State Examination (MMSE) scores, executive function, and episodic memory. A recent study using data from the English Longitudinal Study of Ageing (ELSA) suggested accelerated declines in domains of global cognition, orientation, memory, and executive function after diabetes onset. A key finding of our study lies in the different patterns of cognitive decline. Different from the ELSA study from the UK, the orientation ability in our study did not deteriorate after diabetes (15). We were unable to explore the difference. Future neuroimaging studies could learn the reasons for different cognitive patterns. Moreover, this study firstly report the trends in visuospatial abilities before and after diabetes diagnosis, providing a fuller picture of the cognitive patterns. Our results of the episodic memory and attention and calculation did not meet our threshold of significance, which might be due to different disease duration, the cognitive function tests used, and the length of follow-up. In accordance with the previous two studies, accelerated cognitive decline occurs after but not before diabetes onset. Close monitoring of cognitive dysfunction in patients with diabetes is vitally important in the years following diagnosis.

Another important finding was our subgroup analyses by age. A Whitehall II study pointed out that every 5-year younger age at diabetes onset was associated with a higher dementia risk at age 70 (hazard ratio, 1.24; 95% CI, 1.06 to 1.46) (16). This intrigued us whether younger diabetes patients exhibited more accelerated cognitive decline compared with older patients. However, we did not find age groups modified the effect of diabetes onset on the changes in cognitive slope after diabetes. The previous Whitehall II study collected data on diabetes onset and dementia diagnosis from 1985 to 2019. The follow-up period of CHARLS is relatively short. The current study could be updated after CHALRS released its new survey in the future and explore whether new findings could be achieved after the timeline is prolonged.

Although the exact pathophysiological pathway through which diabetes contributes to cognitive decline beyond aging pathologies remains unclear, several mechanisms have been proposed (25). Increasing evidence suggests that insulin resistance might be a key factor. Brain insulin resistance may lead to impairments in synaptic, metabolic, and immune response functions, which further damage cognitive function (26). Moreover, diabetes induces Alzheimer’s disease-related pathology, such as β-amyloid neurotic plaques (NPs) and intracellular neurofibrillary tangles (NFTs) formed of hyperphosphorylated tau protein (27). Another vital mechanism linking hyperglycemia to cognitive impairment is various vascular damages like stroke, large and small vessel arteriosclerosis, and cerebral amyloid angiopathy. Studies also reported that increased white matter hyperintensities and lacunar infarcts were observed in diabetes patients, which were associated with cognitive deterioration (25, 26, 28, 29). In addition, oxidative stress, neuroinflammation, and some risk factors accompanying diabetes like hypertension, obesity, and depression all contribute to cognition impairment (25, 30, 31, 32, 33). In summary, multiple mechanisms might jointly contribute to brain injury, eventually resulting in post-diabetes cognitive decline. Our previous CHARLS study reported that the stroke patients suffered from an acute decline at stroke onset but not accelerated future decline in global cognition (24). In this study, diabetes patients did not exhibit ‘acute cognitive decline’ in all domains, illustrating that the effect of diabetes-related pathology on the brain tends to be a gradual process, instead of a ‘sudden event’ like stroke. Furthermore, this study was the first to find the slope of visuospatial abilities steeper after diabetes, providing a fuller picture of the cognitive patterns.

Our study has several advantages. This is one of the largest studies exploring the cognitive decline during the before-diabetes period and in the years after new-onset diabetes. To the best of our knowledge, this is the first study learning the cognitive trends among the Chinese population with new-onset diabetes. The large sample size with repeated measurements offered us the opportunity to generate precise and reliable trajectories of cognitive function. Furthermore, our results extend the findings of previous studies by subgroup analyses of younger participants.

A few limitations of this study should be mentioned. First, the CHALRS mainly included Chinese participants in rural areas, limiting the generalizability to other populations. Second, from Wave 1 to Wave 3, the CHARLS lack tests like MMSE. This prevented us from defining dementia or mild cognitive impairment (MCI) by using cut-off points of cognitive scores (34, 35). Among our participants, the cognitive trajectories were similar across different age subgroups. However, we were unable to study whether younger participants had higher risk of dementia, which is a topical research area. Third, CHARLS only performed blood tests in Wave 1 and Wave 3. A certain proportion of diabetes patients were unaware of their glucose level, leading to a delay in diagnosis of diabetes. Future studies could conduct fasting blood tests more frequently. Fourth, also a variety of confounding factors were adjusted, there may still be some related but uncontrolled variables like apolipoprotein E (ApoE) genotype and air pollution. The frequency of the ApoE ε4 allele is about 7% in the Chinese population (36). Hence, the genotype may have fewer effects on our results.

In conclusion, our findings demonstrated that new-onset diabetes was associated with faster rate of future cognitive decline, but not during the pre-diabetes period. Future studies are needed to learn the mechanisms of cognitive decline in a few years after new-onset diabetes.

## Supporting information

STROBE

## Data Availability

Data used in this manuscript from the China Health and Retirement Longitudinal Study (CHARLS). We applied permission for the data access (http://charls.pku.edu.cn/zh-CN) and obtained access to use it. Prof. Yaohui Zhao (National School of Development of Peking University), John Strauss (University of Southern California), and Gonghuan Yang (Chinese Center for Disease Control and Prevention) are the principal investigators. Requests to access these data can also be directed to Jianian Hua.

## List of abbreviations

ApoE: apolipoprotein E
CHARLS: China Health and Retirement Longitudinal Study
ELSA: English Longitudinal Study of Ageing
HbA_1c_: hemoglobin A_1c_
MMSE: Mini-Mental State Examination
NFTs: neurofibrillary tangles
NPs: neurotic plaques
IADLs: instrumental activities of daily living.

## 5 Acknowledgments

We appreciated the China Center for Economic Research and the National School of Development of Peking University for providing the data.

## 7 Supporting information

Checklist S1. STROBE statement.

(DOCX)

Appendix S1. Text S1. Ethic Approval; Fig S1. The conceptual model of our main analysis; Fig S2. The conceptual model of analysis of risk factors; Fig S3. The conceptual model of analysis of acute cognitive decline; Table S1. Number of diabetes onset according to different definitions; Table S2. Number of available cognition measurements in each wave; Table S3. Comparison of baseline characteristics between participants included (n=12422) and excluded due to loss to follow-up (n=1377); Table S4. Trajectories of cognitive z scores among all participants over time; Table S5. Changes in cognitive z scores after diabetes onset according to sex; Table S6. Changes in cognitive z scores after diabetes onset according to education level; Table S7. Changes in cognitive z scores after diabetes onset according to baseline cognitive tertiles; Table S8. Changes in cognitive z scores after diabetes onset according to residential area; Table S9. Changes in cognitive z scores after diabetes onset according to medication after diabetes; Table S10. Trajectories of cognitive z scores among all participants over time after considering acute cognitive decline after diabetes; Table S11. Trajectories of cognitive z scores among all participants over time after adjusting for BMI; Table S12. Trajectories of cognitive z scores among all participants over time after excluding participants experiencing a stroke during the follow-up; Table S13. Trajectories of cognitive z scores among all participants over time only including participants who received cognitive tests in all four waves.

(DOCX)

