## Appendix S1 for "Trends in cognitive function before and after diabetes onset in China"

### 1 Appendix Tests

#### 1.1 Text S1. Ethics approval

Ethical approval for all the CHARLS waves was granted from the Institutional Review Board at Peking University. The IRB approval number for the main household survey, including anthropometrics, is IRB00001052-11015; the IRB approval number for biomarker collection is IRB00001052-11014. During the fieldwork, each respondent who agreed to participate in the survey was asked to sign two copies of informed consent; one copy was kept in the CHARLS office, which was also scanned and saved in PDF format. Four separate consents were obtained: one for the main fieldwork, one for the non-blood biomarkers and one for the taking of the blood samples, and another for storage of blood for future analyses.

#### 1.2 Text S2. Methods

**Covariates**

Educational attainment was divided into illiterate, primary school, middle school, and high school and above. Marital status was dichotomized into married and other status. Body mass index (BMI) was categorized into underweight ($<$18.5 kg/m2), normal weight (18.5–23.9 kg/m2), and overweight or obese (≥24 kg/m2). Hypertension was defined as self-reported diagnosis, use of antihypertensive medication, or the mean of three systolic/diastolic blood pressure measurements ≥140/90 mm Hg. High total cholesterol was defined as self-reported diagnosis, use of lipid-lowering medication, or serum total cholesterol ≥ 240mg/dl. Heart diseases included heart attack, coronary heart disease, angina, congestive heart failure, and other heart problems. Depressive symptoms were measured using the 10-item Centre for Epidemiologic Studies Short Depression Scale, with depression defined as a score of ≥12 from a total score of 0 to 30.

**Statistical analysis**

We constructed a linear mixed model to analyze the longitudinal dataset with repeated measurements. Time is our level-1 variable, and participant is the level-2 variable. We fitted fixed effects for intercept, diabetes (yes or no), time (years since baseline), ‘diabetes*time’ interaction, ‘diabetes*diabetes status*time after diabetes’ interaction, and all the covariates. We fitted random effects for intercept and slope (time since baseline and time after diabetes) to accommodate the correlation of cognitive measures within participants over time, to allow for the variation in average cognitive scores across participants, and to allow for the variation in average cognitive change rate across participants. The fixed effect parameter estimate for the variable ‘diabetes’ indicated whether there were difference in cognitive function between the diabetes group and the control group without diabetes at baseline (time=0). The ‘diabetes*time’ interaction calculated the difference between the average pre-diabetes cognitive change rate for the diabetes group and the average change rate from baseline to the end of follow-up for the control group. The ‘diabetes status’ was a time-varying variable that changed from ‘0’ to ‘1’ at the time of diabetes onset. In this study, the primary coefficient of interest is the ‘diabetes*diabetes status*time after diabetes’ interaction. It reflected changes in cognitive slope before and after diabetes onset among participants who developed diabetes.

### 2 Appendix Figures

#### 2.1 Figure S1. The conceptual model of our main analysis.

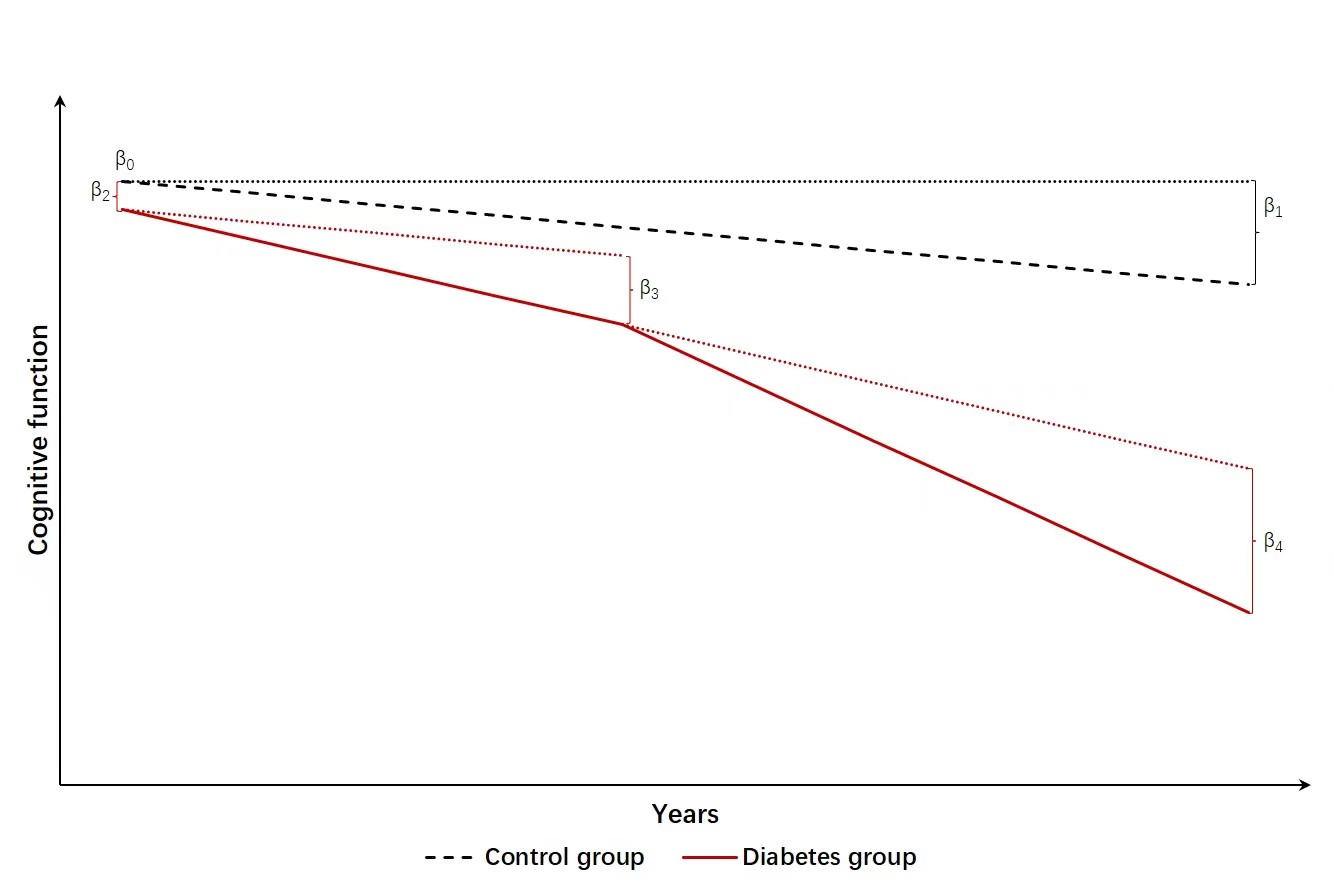

The conceptual model of our study. Time on the x-axis is the years from the date of the first cognitive test. Y-axis is the cognitive function. The black dashed line represents the possible trajectory of the control group without diabetes. We hypothesized their cognitive function declined annually due to aging. The cognitive trajectories of the diabetes group (red lines) consisted of the trajectories before diabetes and the accelerated decline after diabetes.

$\beta$0: The predicted cognition of the without-diabetes group at time t=0.

$\beta$1: The difference in cognition from time t to time t+1 among the without- diabetes group. In other words, it’s the average slope of cognition of the entire without- diabetes group.

$\beta$2: The difference in cognition at time t=0 in the diabetes group compared to the without-diabetes group. It was named ‘baseline difference’ in Table 2.

$\beta$3: The difference in slope in the diabetes group compared to the without-diabetes group in the pre-diabetes period.

$\beta$4: The change in slope in the post-diabetes period compared to the pre-diabetes period. After diabetes, we hypothesized the cognitive decline rate was combined with the pre-diabetes decline rate and an accelerated decline caused by diabetes. We assumed diabetes affects cognition in all years after diabetes.

#### 2.2 Fig S2. The conceptual model of analysis of risk factors

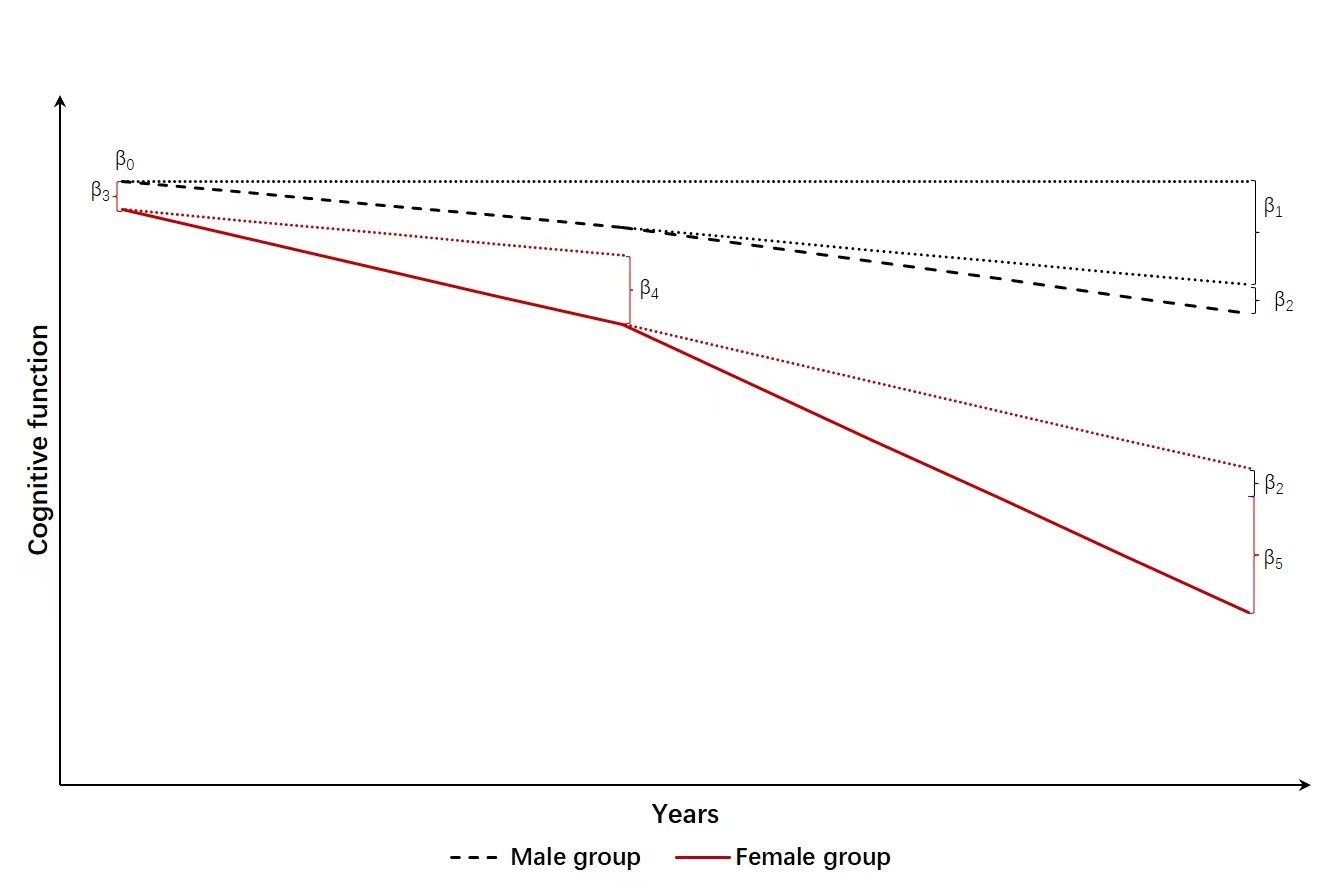

This analysis calculated the effect of risk factors (female sex) on the effect of diabetes on cognitive trajectories. Time on the x-axis is the years from the date of the first cognitive test. Y-axis is the cognitive function. The black dashed line represents the possible trajectory of the male participants (reference group) with diabetes onset. While the red solid line represents the possible trajectory of the female participants with diabetes onset.

$\beta$0: The predicted cognition of the male group at time t=0.

$\beta$1: The difference in cognition from time t to time t+1 among the male group.

$\beta$2: The change in slope in the post-diabetes period compared to the pre-diabetes period among the male group.

$\beta$3: The difference in cognition at time t=0 in the female group compared to the male group.

$\beta$4: The difference in slope in the female group compared to the male group in the pre-diabetes period.

$\beta$5: The effect of female sex on the effect of diabetes on the changes in cognitive slope after diabetes onset.

#### 2.3 Fig S3. The conceptual model of analysis of acute cognitive decline

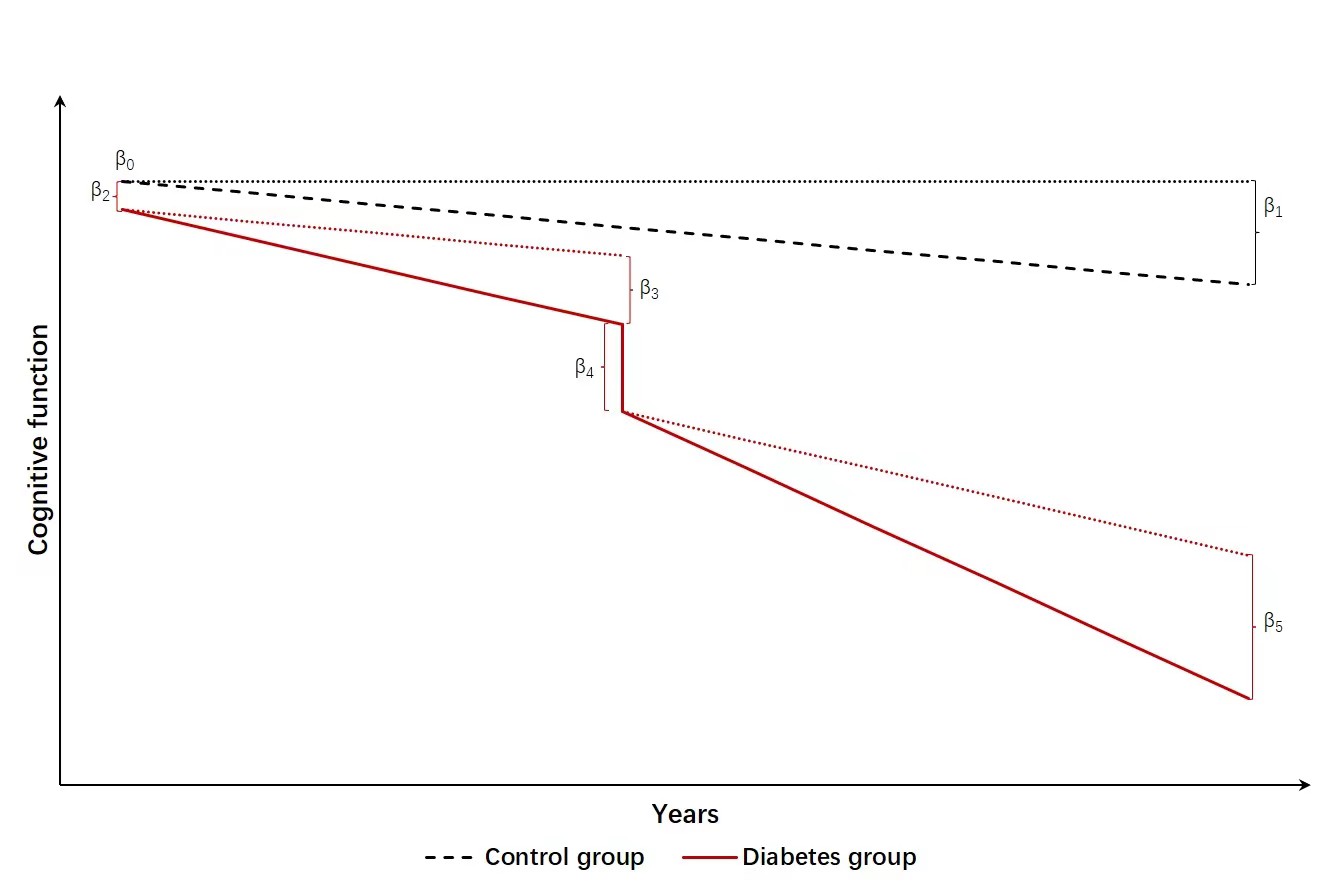
This model was based on Figure S1 and added an ‘acute cognitive decline’ represented by $\beta$4.

$\beta$4: The ‘acute cognitive change’ at the time of diabetes among the diabetes group, measured as the first post- diabetes cognitive score minus the last pre- diabetes cognitive score.

Let us hypothesize a person received cognitive tests in 2011, 2013, 2015, and 2018, and was diagnosed with diabetes in 2014. We define C_2011_ as the cognitive score in 2011. C_2015_ $-$ C_2013_ $=$ (C_2014_ $-$ C_2013_) $+$ (C_2015_ $-$ C_2014_) *we don’t know the cognitive score in 2014, since the score wasn’t recorded during this year* = annual decline rate before diabetes + annual decline rate after diabetes $=$ (C_2013_ $-$ C_2011_)^*^1/2 $+$ (C_2018_ $-$ C_2015_)^*^1/3. But, the actual C_2015_ $-$ C_2013_ is more than (C_2013_ $-$ C_2011_)^*^1/2 $+$ (C_2018_ $-$ C_2015_)^*^1/3, which means the cognitive scores declined more than expected. The decline exceeding expectation is the “acute cognitive decline”.

### 3 Appendix Tables

#### 3.1 Table S1. Number of diabetes onset according to different definitions

| Definition | Diagnosis time | Number of new-onset diabetes |
| --- | --- | --- |
| Self-reported ($+$)^a^ |  |  |
|  | 0 to 2 years | 259 |
|  | 2 to 4 years | 106 |
|  | 4 to 7 years | 356 |
| Self-reported ($-$) Medication ($+$)^b^ | | |
|  | 0 to 2 years | 0 |
|  | 2 to 4 years | 68 |
|  | 4 to 7 years | 64 ( |
| Self-reported ($-$) Medication ($-$) HbA_1c_ ($+$) | | |
|  | 0 year | 64^c^ |
|  | 0 to 4 years | 353 |

^a^Reported diagnosis of diabetes by a doctor.

^b^Did no report diagnosis of diabetes but reported use of insulin or antihyperglycemic medication.

^c^Did no report diagnosis of diabetes, insulin, or antihyperglycemic medication. But HbA_1c_ $\geq$6.5%. 64 participants were excluded from our study due to baseline diabetes.

#### 3.2 Table S2. Number of available cognition measurements in each wave

|  | Wave 1 | Wave 2 | Wave 3 | Wave 4 |
| --- | --- | --- | --- | --- |
| No-diabetes | 11215 (100%) | 9974 (88.9%) | 9675 (86.3%) | 7678 (68.5%) |
| Diabetes | 1207 (100%) | 1112 (92.1%) | 1120 (92.8%) | 930 (77.1%) |

#### 3.3 Table S3. Comparison of baseline characteristics between participants included (n=12422) and excluded due to loss to follow-up (n=1377)^a^

|  | Included | Excluded | *p* value^a^ |
| --- | --- | --- | --- |
|  | (n = 12422) | (n = 1377) |  |
| Continuous variables, mean (SD) |  | | |
| Age | 58.6 (9.2) | 64.2 (12.6) | <0.001 |
| Number of IADLs | 0.2 (0.6) | 0.5 (1.2) | <0.001 |
| Episodic memory | 3.3 (1.9) | 2.7 (2.2) | <0.001 |
| Visuospatial abilities | 0.6 (0.5) | 0.5 (0.5) | <0.001 |
| Orientation | 3.7 (1.4) | 3.5 (1.7) | <0.001 |
| Calculation and attention | 2.8 (2.0) | 2.5 (2.1) | <0.001 |
| Categorical variables, n (%) |  | | |
| Males | 5911 (47.4) | 624 (49.3) | 0.197 |
| Education |  |  | <0.001 |
| Illiterate | 3347 (26.9) | 419 (33.3) |  |
| Primary school | 5000 (40.1) | 413 (32.7) |  |
| Middle school | 2617 (21.0) | 221 (17.5) |  |
| High school and above | 1503 (12.1) | 208 (16.5) |  |
| Marital status |  |  | <0.001 |
| Married | 11070 (88.7) | 982 (77.7) |  |
| Other status | 1402 (11.3) | 283 (22.3) |  |
| Residential area |  |  | <0.001 |
| Urban | 2250 (20.2) | 536 (42.4) |  |
| Rural | 9933 (79.8) | 727 (57.6) |  |
| Current smoking | 4883 (39.2) | 521 (41.2) | 0.158 |
| Current drinking | 3221 (25.8) | 277 (21.9) | 0.002 |
| Hypertension | 3483 (28.0) | 439 (35.1) | <0.001 |
| High total cholesterol | 1733 (14.0) | 145 (11.6) | 0.020 |
| Cancer | 104 (0.8) | 22 (1.7) | 0.003 |
| Lung Diseases | 1196 (9.6) | 156 (12.3) | 0.002 |
| Heart problems | 1316 (10.6) | 168 (13.3) | 0.003 |
| Depression | 2575 (20.7) | 279 (22.1) | 0.239 |

^a^Calculated using ANOVA for continuous covariates and χ^2^ test for categorical covariates.

#### 3.4 Table S4. Trajectories of cognitive z scores among all participants over time^a^

|  | Global cognition |  | Episodic Memory |  | Visuospatial ability |  | Orientation |  | Attention and calculation |  |
| --- | --- | --- | --- | --- | --- | --- | --- | --- | --- | --- |
|  | β (95% CI) | *p* | β (95% CI) | *p* | β (95% CI) | *p* | β (95% CI) | *p* | β (95% CI) | *p* |
| Variables^b^ |  |  |  |  |  |  |  |  |  |  |
| Baseline age | -0.017 (-0.018, -0.016) | <0.001 | -0.023 (-0.024, -0.021) | <0.001 | -0.012 (-0.013, -0.010) | <0.001 | -0.009 (-0.010, -0.008) | <0.001 | -0.006 (-0.007, -0.005) | <0.001 |
| Intercept of without-diabetes group | 0.136 (0.023, 0.248) | 0.018 | 0.392 (0.277, 0.507) | <0.001 | 0.198 (0.088, 0.308) | <0.001 | 0.070 (-0.035, 0.176) | 0.190 | 0.177 (0.063, 0.291) | 0.002 |
| Difference in baseline | -0.006 (-0.055, 0.042) | 0.802 | 0.018 (-0.039, 0.076) | 0.530 | -0.025 (-0.082, 0.033) | 0·4004 | -0.009 (-0.057, 0.039) | 0.713 | -0.019 (-0.074, 0.036) | 0.496 |
| Slope of without-diabetes group | -0.035 (-0.038, -0.032) | <0.001 | -0.005 (-0.008, -0.001) | 0.005 | -0.037 (-0.041, -0.034) | <0.001 | -0.035 (-0.038, -0.033) | <0.001 | -0.053 (-0.055, -0.050) | <0.001 |
| Difference in slope before diabetes | 0.006 ( -0.007, 0.019) | 0.376 | <0.001 (-0.016, 0.015) | 0.951 | 0.011 (-0.005, 0.028) | 0.178 | 0.003 (-0.009, 0.016) | 0.603 | 0.010 (-0.004, 0.024) | 0.174 |
| Changes in slope after diabetes | -0.023 (-0.043, -0.004) | 0.019 | -0.018 (-0.041, 0.004) | 0.116 | -0.036 (-0.061, -0.011) | 0.004 | 0.001 (-0.018, 0.020) | 0.894 | -0.017 (-0.037, 0.003) | 0.090 |
| -2log likelihood | 86423.6 |  | 105440.4 |  | 108927.6 |  | 103949.6 |  | 111780.9 |  |

^a^Adjusted for baseline age, sex, education, marital status, residential area, current smoking, current drinking, hypertension, dyslipidaemia, cancer, lung diseases, heart problems, depression, and number of IADLs.

^b^Detailed description for variables is shown in Figure S1.

#### 3.5 Table S5. Changes in cognitive z scores after diabetes onset according to sex^a^

|  | Male | Female | *p* for interaction^b^ |
| --- | --- | --- | --- |
| Global cognition | -0.035 (-0.065, -0.006) | -0.016 (-0.041, 0.010) | 0.288 |
| Episodic memory | -0.026 (-0.061, 0.009) | -0.016 (-0.045, 0.013) | 0.699 |
| Visuospatial abilities | -0.056 (-0.093, -0.019) | -0.024 (-0.057, 0.009) | 0.227 |
| Orientation | 0.010 (-0.021, 0.041) | -0.002 (-0.027, 0.023) | 0.589 |
| Attention and calculation | -0.019 (-0.054, 0.016) | -0.017 (-0.042, 0.007) | 0.934 |

^a^Adjusted for baseline age, education, marital status, residential area, current smoking, current drinking, hypertension, dyslipidaemia, cancer, lung diseases, heart problems, depression, and number of IADLs.

^b^The interaction term reflected whether age groups modified the effect of diabetes on the cognitive decline rate after diabetes onset.

#### 3.6 Table S6. Changes in cognitive z scores after diabetes onset according to education level^a^

|  | Illiterate and primary school | Middle school | High school and above |
| --- | --- | --- | --- |
| Global cognition |  |  |  |
| β (95% CI) | -0.040 (-0.078, -0.003) | -0.015 (-0.047, 0.018) | -0.023 (-0.053, 0.008) |
| *p* for interaction^b^ | Ref. | 0.219 | 0.415 |
| Episodic memory |  |  |  |
| β (95% CI) | -0.058 (-0.096, -0.020) | 0.001 (-0.037, 0.039) | -0.009 (-0.047, 0.028) |
| *p* for interaction | Ref. | 0.012 | <0.001 |
| Visuospatial abilities |  |  |  |
| β (95% CI) | -0.026 (-0.073, 0.021) | -0.024 (-0.066, 0.018) | -0.045 (-0.083, -0.008) |
| *p* for interaction | Ref, | 0.924 | 0.467 |
| Orientation |  |  |  |
| β (95% CI) | -0.014 (-0.05, 0.023) | 0.012 (-0.020, 0.044) | 0.002 (-0.029, 0.034) |
| *p* for interaction | Ref. | 0.280 | 0.376 |
| Attention and calculation |  |  |  |
| β (95% CI) | -0.032 (-0.06, -0.004) | -0.015 (-0.049, 0.019) | -0.009 (-0.049, 0.031) |
| *p* for interaction | Ref. | 0.341 | 0.225 |

^a^Adjusted for baseline age, sex, marital status, residential area, current smoking, current drinking, hypertension, dyslipidaemia, cancer, lung diseases, heart problems, depression, and number of IADLs.

^b^The interaction term reflected whether age groups modified the effect of diabetes on the cognitive decline rate after diabetes onset.

#### 3.7 Table S7. Changes in cognitive z scores after diabetes onset according to baseline cognitive tertiles^a^

|  | Lowest tertile | Median tertile | Highest tertile |
| --- | --- | --- | --- |
| Global cognition |  |  |  |
| β (95% CI) | -0.097 (-0.132, -0.061) | -0.012 (-0.055, 0.031) | 0.041 (0.009, 0.074) |
| *p* for interaction^b^ | Ref. | <0.001 | <0.001 |
| Episodic memory |  |  |  |
| β (95% CI) | -0.159 (-0.208, -0.109) | -0.012 (-0.055, 0.031) | 0.066 (0.03, 0.102) |
| *p* for interaction | Ref. | <0.001 | <0.001 |
| Orientation |  |  |  |
| β (95% CI) | -0.106 (-0.162, -0.049) | None | 0.019 (-0.002, 0.04) |
| *p* for interaction | Ref. | None | <0.001 |
| Attention and calculation |  |  |  |
| β (95% CI) | -0.121 (-0.151, -0.09) | -0.041 (-0.089, 0.007) | 0.105 (0.056, 0.154) |
| *p* for interaction | Ref. | <0.001 | <0.001 |

^a^Adjusted for baseline age, sex, education, marital status, residential area, current smoking, current drinking, hypertension, dyslipidaemia, cancer, lung diseases, heart problems, depression, and number of IADLs.

^b^The interaction term reflected whether age groups modified the effect of diabetes on the cognitive decline rate after diabetes onset.

#### 3.8 Table S8. Changes in cognitive z scores after diabetes onset according to residential area^a^

|  | Rural areas | Urban areas | *p* for interaction^b^ |
| --- | --- | --- | --- |
| Global cognition | -0.021 (-0.044, 0.001) | -0.026 (-0.065, 0.012) | 0.820 |
| Episodic memory | -0.026 (-0.052, -0.001) | 0.007 (-0.040, 0.054) | 0.183 |
| Visuospatial abilities | -0.028 (-0.056, <0.001) | -0.055 (-0.103, -0.006) | 0.345 |
| Orientation | 0.011 (-0.011, 0.033) | -0.033 (-0.075, 0.008) | 0.105 |
| Attention and calculation | -0.019 (-0.042, 0.003) | -0.004 (-0.054, 0.045) | 0.664 |

^a^Adjusted for baseline age, sex, education, marital status, current smoking, current drinking, hypertension, dyslipidaemia, cancer, lung diseases, heart problems, depression, and number of IADLs.

^b^The interaction term reflected whether age groups modified the effect of diabetes on the cognitive decline rate after diabetes onset.

#### 3.9 Table S9. Changes in cognitive z scores after diabetes onset according to medication after diabetes^a^

|  | Using antidiabetic medication | Without antidiabetic medication | *p* for interaction^b^ |
| --- | --- | --- | --- |
| Global cognition | -0.044 (-0.075, -0.013) | -0.007 (-0.031, 0.018) | 0.037 |
| Episodic memory | -0.018 (-0.052, 0.016) | -0.017 (-0.047, 0.012) | 0.231 |
| Visuospatial abilities | -0.047 (-0.087, -0.008) | -0.023 (-0.055, 0.008) | 0.263 |
| Orientation | -0.007 (-0.040, 0.025) | 0.008 (-0.017, 0.033) | 0.496 |
| Attention and calculation | -0.042 (-0.075, -0.009) | -0.002 (-0.028, 0.024) | 0.061 |

^a^Adjusted for baseline age, sex, education, marital status, residential area, current smoking, current drinking, hypertension, dyslipidaemia, cancer, lung diseases, heart problems, depression, and number of IADLs.

^b^The interaction term reflected whether age groups modified the effect of diabetes on the cognitive decline rate after diabetes onset.

#### 3.10 Table S10. Trajectories of cognitive z scores among all participants over time after considering acute cognitive decline after diabtes^ab^

|  | Global cognition |  | Episodic Memory |  | Visuospatial ability |  | Orientation |  | Attention and calculation |  |
| --- | --- | --- | --- | --- | --- | --- | --- | --- | --- | --- |
|  | β (95% CI) | *p* | β (95% CI) | *p* | β (95% CI) | *p* | β (95% CI) | *p* | β (95% CI) | *p* |
| Variables^b^ |  |  |  |  |  |  |  |  |  |  |
| Baseline age | -0.017 (-0.018, -0.016) | <0.001 | -0.023 (-0.024, -0.021) | <0.001 | -0.012 (-0.013, -0.010) | <0.001 | -0.009 (-0.010, -0.008) | <0.001 | -0.006 (-0.007, -0.005) | <0.001 |
| Intercept of without-diabetes group | 0.136 (0.023, 0.248) | 0.018 | 0.391 (0.276, 0.507) | <0.001 | 0.198 (0.089, 0.308) | <0.001 | 0.071 (-0.034, 0.176) | 0.187 | 0.177 (0.063, 0.291) | 0.002 |
| Difference in baseline | -0.006 (-0.055, 0.043) | 0.823 | 0.014 (-0.044, 0.072) | 0.631 | -0.024 (-0.081, 0.034) | 0.423 | -0.004 (-0.053, 0.044) | 0.857 | -0.019 (-0.074, 0.036) | 0.503 |
| Slope of without-diabetes group | -0.035 (-0.038, -0.032) | <0.001 | -0.005 (-0.008, -0.001) | 0.005 | -0.037 (-0.041, -0.034) | <0.001 | -0.035 (-0.038, -0.033) | <0.001 | -0.053 (-0.055, -0.050) | <0.001 |
| Difference in slope before diabetes | 0.005 (-0.009, 0.020) | 0.493 | 0.005 (-0.013, 0.023) | 0.582 | 0.010 (-0.009, 0.028) | 0.291 | -0.002 (-0.017, 0.013) | 0.821 | 0.010 (-0.007, 0.026) | 0.253 |
| Acute cognitive decline | 0.007 (-0.055, 0.069) | 0.828 | -0.054 (-0.130, 0.023) | 0.168 | 0.015 (-0.071, 0.100) | 0.737 | 0.045 (-0.024, 0.114) | 0.203 | 0.001 (-0.073, 0.076) | 0.973 |
| Changes in slope after diabetes | -0.024 (-0.044, -0.004) | 0.020 | -0.013 (-0.037, 0.011) | 0.275 | -0.037 (-0.064, -0.011) | 0.005 | -0.002 (-0.021, 0.018) | 0.879 | -0.017 (-0.038, 0.003) | 0.098 |
| -2log likelihood | 86423.6 |  | 105438.5 |  | 108927.4 |  | 103948.0 |  | 111780.9 |  |

^a^Adjusted for baseline age, sex, education, marital status, residential area, current smoking, current drinking, hypertension, dyslipidaemia, cancer, lung diseases, heart problems, depression, and number of IADLs.

^b^Detailed description for variables is shown in Figure S3.

#### 3.11 Table S11 Trajectories of cognitive z scores among all participants over time after adjusting for BMI^a^

|  | Global cognition |  | Episodic Memory |  | Visuospatial ability |  | Orientation |  | Attention and calculation |  |
| --- | --- | --- | --- | --- | --- | --- | --- | --- | --- | --- |
|  | β (95% CI) | *p* | β (95% CI) | *p* | β (95% CI) | *p* | β (95% CI) | *p* | β (95% CI) | *p* |
| Variables^b^ |  |  |  |  |  |  |  |  |  |  |
| Baseline age | -0.017 (-0.019, -0.016) | <0.001 | -0.023 (-0.025, -0.022) | <0.001 | -0.012 (-0.013, -0.010) | <0.001 | -0.010 (-0.011, -0.008) | <0.001 | -0.007 (-0.009, -0.006) | <0.001 |
| Intercept of without-diabetes group | -0.150 (-0.309, 0.009) | 0.065 | 0.233 (0.074, 0.392) | 0.004 | 0.022 (-0.132, 0.177) | 0.775 | -0.139 (-0.286, 0.008) | 0.064 | 0.075 (-0.085, 0.235) | 0.358 |
| Difference in baseline | -0.012 (-0.066, 0.041) | 0.656 | 0.029 (-0.033, 0.092) | 0.360 | -0.034 (-0.097, 0.030) | 0.296 | -0.029 (-0.081, 0.023) | 0.274 | -0.022 (-0.082, 0.037) | 0.464 |
| Slope of without-diabetes group | -0.037 (-0.040, -0.034) | <0.001 | -0.008 (-0.012, -0.004) | <0.001 | -0.039 (-0.043, -0.035) | <0.001 | -0.038 (-0.041, -0.035) | <0.001 | -0.054 (-0.057, -0.050) | <0.001 |
| Difference in slope before diabetes | 0.004 (-0.010, 0.019) | 0.539 | -0.005 (-0.022, 0.012) | 0.556 | 0.012 (-0.007, 0.030) | 0.216 | 0.005 (-0.009, 0.018) | 0.524 | 0.011 (-0.004, 0.026) | 0.151 |
| Changes in slope after diabetes | -0.022 (-0.043, -0.001) | 0.038 | -0.018 (-0.043, 0.006) | 0.149 | -0.037 (-0.064, -0.010) | 0.007 | 0.004 (-0.016, 0.024) | 0.710 | -0.017 (-0.038, 0.005) | 0.131 |
| -2log likelihood | 69465.7 |  | 64326.3 |  | 87989.2 |  | 81512.9 |  | 88870.8 |  |

^a^Adjusted for baseline age, sex, education, marital status, residential area, current smoking, current drinking, hypertension, dyslipidaemia, cancer, lung diseases, heart problems, depression, and number of IADLs.

^b^Detailed description for variables is shown in Figure S1.

#### 3.12 Table S12. Trajectories of cognitive z scores among all participants over time after excluding participants experiencing a stroke during the follow-up^a^

|  | Global cognition |  | Episodic Memory |  | Visuospatial ability |  | Orientation |  | Attention and calculation |  |
| --- | --- | --- | --- | --- | --- | --- | --- | --- | --- | --- |
|  | β (95% CI) | *p* | β (95% CI) | *p* | β (95% CI) | *p* | β (95% CI) | *p* | β (95% CI) | *p* |
| Variables^b^ |  |  |  |  |  |  |  |  |  |  |
| Baseline age | -0.017 (-0.018, -0.016) | <0.001 | -0.023 (-0.024, -0.022) | <0.001 | -0.012 (-0.013, -0.010) | <0.001 | -0.009 (-0.010, -0.008) | <0.001 | -0.006 (-0.007, -0.005) | <0.001 |
| Intercept of without-diabetes group | 0.151 (0.035, 0.266) | 0.011 | 0.404 (0.285, 0.523) | <0.001 | 0.201 (0.088, 0.314) | 0.001 | 0.092 (-0.016, 0.200) | 0.095 | 0.180 (0.063, 0.278) | 0.003 |
| Difference in baseline | -0.018 (-0.069, 0.034) | 0.498 | 0.027 (-0.034, 0.088) | 0.381 | -0.039 (-0.099, 0.022) | 0.210 | -0.016 (-0.066, 0.035) | 0.543 | -0.030 (-0.088, 0.027) | 0.300 |
| Slope of without-diabetes group | -0.034 (-0.037, -0.032) | <0.001 | -0.004 (-0.007, -0.001) | 0.024 | -0.037 (-0.040, -0.033) | <0.001 | -0.035 (-0.037, -0.032) | <0.001 | -0.052 (-0.054, -0.049) | <0.01 |
| Difference in slope before diabetes | 0.012 (-0.002, 0.025) | 0.090 | <0.001 (-0.017, 0.016) | 0.962 | 0.015 (-0.002, 0.033) | 0.088 | 0.006 (-0.007, 0.020) | 0.370 | 0.016 (0.001, 0.031) | 0.032 |
| Changes in slope after diabetes | -0.029 (-0.049, -0.008) | 0.006 | -0.018 (-0.042, 0.007) | 0.156 | -0.039 (-0.065, -0.013) | 0.003 | -0.002 (-0.022, 0.018) | 0.862 | -0.025 (-0.046, -0.003) | 0.022 |
| -2log likelihood | 81783.4 |  | 99970.1 |  | 103116.5 |  | 98102.1 |  |  |  |

^a^Adjusted for baseline age, sex, education, marital status, residential area, current smoking, current drinking, hypertension, dyslipidaemia, cancer, lung diseases, heart problems, depression, and number of IADLs.

^b^Detailed description for variables is shown in Figure S1.

#### 3.13 Table S13. Trajectories of cognitive z scores among all participants over time only including participants who received cognitive tests in all four waves^a^

|  | Global cognition |  | Episodic Memory |  | Visuospatial ability |  | Orientation |  | Attention and calculation |  |
| --- | --- | --- | --- | --- | --- | --- | --- | --- | --- | --- |
|  | β (95% CI) | *p* | β (95% CI) | *p* | β (95% CI) | *p* | β (95% CI) | *p* | β (95% CI) | *p* |
| Variables^b^ |  |  |  |  |  |  |  |  |  |  |
| Baseline age | -0.012 (-0.014, -0.010) | <0.001 | -0.021 (-0.023, -0.019) | <0.001 | -0.007 (-0.009, -0.005) | <0.001 | -0.008 (-0.009, -0.006) | <0.001 | -0.006 (-0.007, -0.004) | <0.001 |
| Intercept of without-diabetes group | -0.037 (-0.185, 0.110) | 0.620 | 0.300 (0.150, 0.451) | <0.001 | 0.152 (0.010, 0.294) | 0.036 | -0.008 (-0.130, 0.115) | 0.904 | 0.135 (0.003, 0.266) | 0.045 |
| Difference in baseline | -0.026 (-0.082, 0.031) | 0.379 | 0.030 (-0.037, 0.098) | 0.379 | -0.055 (-0.124, 0.014) | 0.119 | 0.002 (-0.050, 0.053) | 0.948 | -0.026 (-0.085, 0.032) | 0.373 |
| Slope of without-diabetes group | -0.034 (-0.037, -0.031) | <0.001 | -0.002 (-0.006, 0.002) | 0.338 | -0.043 (-0.047, -0.039) | <0.001 | -0.034 (-0.036, -0.031) | <0.001 | -0.052 (-0.056, -0.049) | <0.001 |
| Difference in slope before diabetes | 0.009 (-0.006, 0.023) | 0.241 | -0.001 (-0.019, 0.017) | 0.907 | 0.017 (-0.003, 0.036) | 0.088 | -0.001 (-0.014, 0.012) | 0.888 | 0.011 (-0.004, 0.026) | 0.139 |
| Changes in slope after diabetes | -0.025 (-0.047, -0.004) | 0.020 | -0.023 (-0.048, 0.003) | 0.079 | -0.036 (-0.064, -0.008) | 0.011 | 0.006 (-0.014, 0.025) | 0.554 | -0.019 (-0.040, 0.001) | 0.069 |
| -2log likelihood | 56704.9 |  | 70257.7 |  | 71882.2 |  | 85360.8 |  | 91919.1 |  |

^a^Adjusted for baseline age, sex, education, marital status, residential area, current smoking, current drinking, hypertension, dyslipidaemia, cancer, lung diseases, heart problems, depression, and number of IADLs.

^b^Detailed description for variables is shown in Figure S1.
